# The role of prisons in disseminating tuberculosis in Brazil: a genomic epidemiology study

**DOI:** 10.1101/2021.06.22.21259360

**Authors:** Katharine S. Walter, Paulo César Pereira dos Santos, Thais Oliveira Gonçalves, Brunna Oliveira da Silva, Andrea da Silva Santos, Alessandra de Cássia Leite, Alessandra Moura da Silva, Flora Martinez Figueira Moreira, Roberto Dias de Oliveira, Everton Ferreira Lemos, Eunice Cunha, Yiran E. Liu, Albert Ko, Caroline Colijn, Ted Cohen, Barun Mathema, Julio Croda, Jason R. Andrews

**Affiliations:** Division of Infectious Diseases and Geographic Medicine, Stanford University School of Medicine; Stanford, United States; Health Sciences Research Laboratory, Federal University of Grande Dourados; Dourados, Brazil; Laboratory of Bacteriology, Central Laboratory of Mato Grosso do Sul; Campo Grande, Brazil; School of Medicine, Federal University of Mato Grosso do Sul, School of Medicine; Campo Grande, Brazil; Nursing Course, State University of Mato Grosso do Sul; Dourados, Brazil; Cancer Biology Graduate Program, Stanford University School of Medicine; Stanford, United States; Department of Epidemiology of Microbial Diseases, Yale School of Public Health; New Haven, United States; Department of Mathematics, Simon Fraser University; Burnaby, Canada; Department of Epidemiology, Columbia University Mailman School of Public Health; New York, United States; Mato Grosso do Sul Office, Oswaldo Cruz Foundation; Campo Grande, Brazil

**Author notes:** Corresponding Author (KSW). These authors contributed equally to this work (Julio Croda, Jason R. Andrews).

**Keywords:** tuberculosis, epidemiology, genomics, disease reservoir, spillover, prisons

## Abstract

**Background:** Globally, prisons are high-incidence settings for tuberculosis. Yet the role of prisons as reservoirs of *M. tuberculosis*, propagating epidemics through spillover from prisons to surrounding communities, has been difficult to measure directly.

**Methods and Findings:** To quantify the role of prisons in driving wider community *M. tuberculosis* transmission, we conducted prospective genomic surveillance in Central West Brazil from 2014 to 2019. We whole genome sequenced 1,152 *M. tuberculosis* isolates collected during active and passive surveillance inside and outside prisons and linked genomes to detailed incarceration histories. We applied multiple phylogenetic and genomic clustering approaches and inferred timed transmission trees. We found that *M. tuberculosis* sequences from incarcerated and non-incarcerated people were closely related in a maximum likelihood phylogeny. The majority (70.8%; 46/65) of genomic clusters including people with no incarceration history also included individuals with a recent history of incarceration. Among cases in individuals with no incarceration history, 50.6% (162/320) were in clusters that included individuals with recent incarceration history, suggesting that transmission chains often span prisons and communities. We identified a minimum of 18 highly probable spillover events, *M. tuberculosis* transmission from people with a recent incarceration history to people with no prior history of incarceration, occurring in the state’s four largest cities and across sampling years. We additionally found that frequent transfers of people between the state’s prisons creates a highly connected prison network that likely disseminates *M. tuberculosis* across the state.

**Conclusions:** We developed a framework for measuring spillover from high-incidence environments to surrounding communities by integrating genomic and spatial information to reconstruct tuberculosis transmission chains. Our findings indicate that, in this setting, prisons serve not only as disease reservoirs, but also disseminate *M. tuberculosis* across highly connected prison networks, both amplifying and propagating *M. tuberculosis* risk in surrounding communities.

## Introduction

Tuberculosis remains one of the leading causes of death by an infectious disease and led to 1.4 million deaths in 2019.[1] Locating where *M. tuberculosis* transmission occurs is critical for developing public health interventions to block transmission and prevent future infections. Yet in high-incidence settings, where most tuberculosis is transmitted remains unknown.[2–7] Built environments often shape *M. tuberculosis* transmission risk, and high-transmission settings like prisons, hospitals, and mines may have epidemiological impacts that transcend their walls[8–10], serving as institutional amplifiers of infection.[11,12] In particular, the conditions of detention, including severe overcrowding, poor ventilation, poor nutrition, and limited access to healthcare, put incarcerated people at elevated risk of many infectious diseases, including tuberculosis.[8–10]

Globally, incidence of tuberculosis in incarcerated populations is more than ten times that in the general population.[9,13] The role of prisons in driving community tuberculosis epidemics is of particular concern in Central and South America, where the incarcerated population has increased by over 200% since 2000 and tuberculosis cases among the incarcerated population have increased by over 269%[14]. Alarmingly, increasing tuberculosis notifications within prisons more than offset declines in tuberculosis outside prisons across the region[14].

The full impact of increasing incarceration rates on the tuberculosis epidemic in the Americas and elsewhere is underestimated by total incident cases within prisons. Many people are infected in prison, but only diagnosed later.[12,15,16] Further, spillover may occur when incarcerated people, prison staff, or visitors are infected within high-incidence prison environments and transmit *M. tuberculosis* onwards in communities outside prison. Previous studies have found that the excess risk of tuberculosis within prisons extends to surrounding communities[17,18] and that *M. tuberculosis* genotypes responsible for jail and prison outbreaks are also found in the surrounding communities[15,17,19,20]. Because transmission is rarely observed, however, it is often difficult to link community cases to transmission from prisons and previous studies have largely been associative.

Pathogen genomes can be powerfully harnessed to infer high-resolution evolutionary and transmission histories, including who-infected-whom.[21] One previous study in Georgia used *M. tuberculosis* genomes to identify frequent transmission of multi-drug resistant tuberculosis from prisons to surrounding communities.[22] However, studies have not yet combined *M. tuberculosis* genomic, epidemiologic, and location data to reconstruct transmission linkages between potential institutional amplifiers and the community. To quantify the contribution of prison spillover to tuberculosis transmission in the community, we conducted prospective genomic surveillance for tuberculosis in the Central West Brazilian state of Mato Grosso do Sul. We integrated *M. tuberculosis* genomes with detailed incarceration histories and took multiple phylogenetic and genomic clustering approaches to reconstruct spillover events, transmission of tuberculosis from prisons to the community.

## Methods

### Study design

We conducted population-based tuberculosis surveillance in the state of Mato Grosso do Sul in Central West Brazil from January 2014 through May 2019. Surveillance included active screening in three of the largest prisons in the state as well as ongoing passive surveillance focused on the two largest cities in the state, and three cities at the state’s border with Paraguay and Bolivia (Fig. 1a,b; **S1 Text**). All participants provided written consent, and this study was conducted with the approval of the Research Ethics Committee from the Federal University of Grande Dourados, Federal University of Mato Grosso do Sul and National Research Ethics Committee (CONEP) (CAAE 37237814.4.0000.5160, 2676613.3.1001.5160, and 26620619.6.0000.0021) and Stanford University Institutional Review Board (IRB-40285) (**S1 Text**).

**Fig 1.**
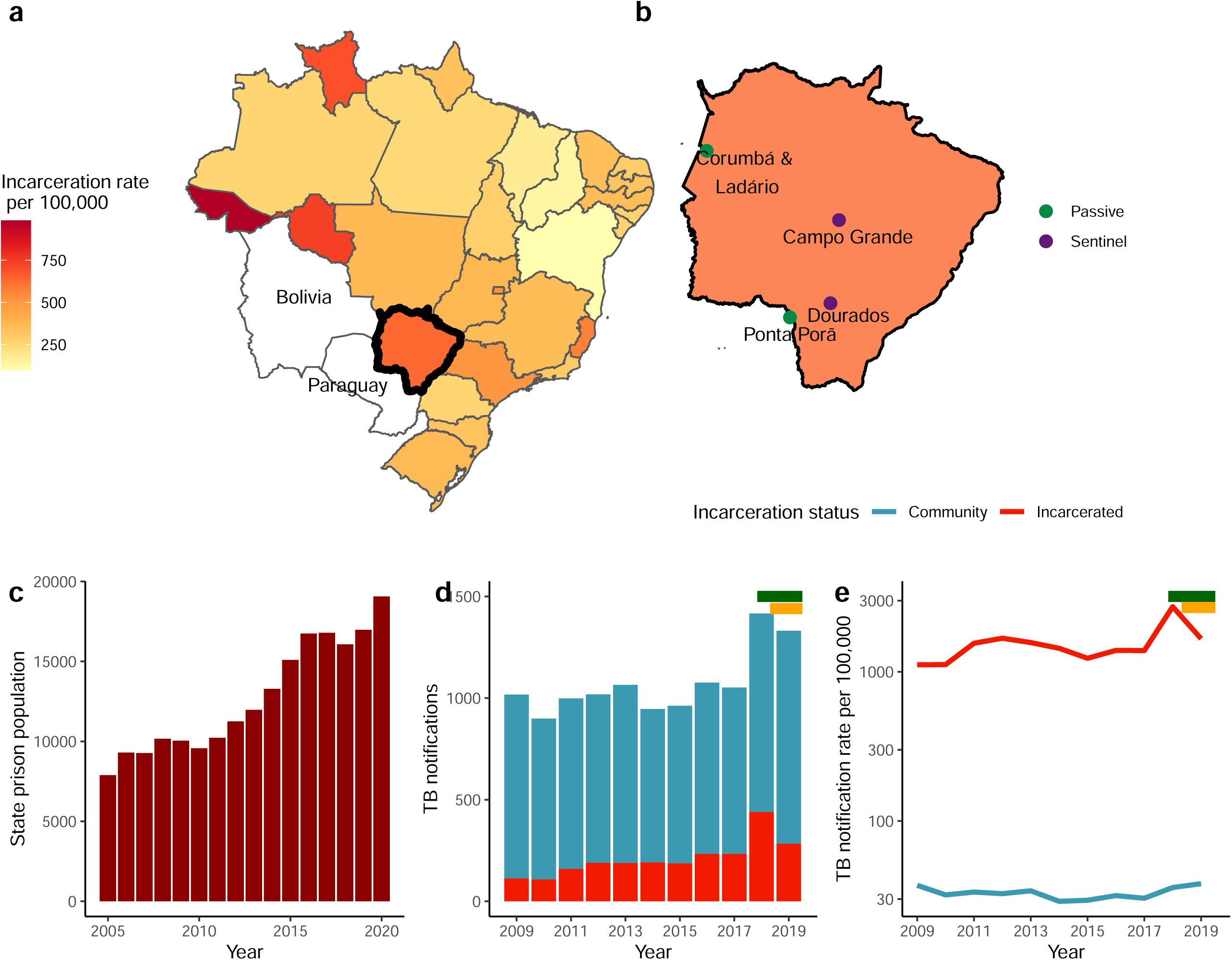
Tuberculosis is increasingly concentrated within prisons in Mato Grosso do Sul, Brazil. (a) Map of Brazil with states colored by the 2019 incarceration rate with Mato Grosso do Sul outlined in black. (b) Map of Mato Grosso do Sul state with the cities with passive surveillance only and where active surveillance was additionally conducted within prisons. (c) The incarcerated population size in Mato Grosso do Sul grew by 142% from 2005 to 2020, increasing from 7,891 to 19,065 people. (d) Mato Grosso do Sul state’s annual new and retreatment tuberculosis notifications, colored by incarceration status and (e) the notification rate per 100,000 people for the incarcerated population and non-incarcerated populations from 2009 to 2019. The y-axis is log-scaled. The mean tuberculosis notification rate was more than 46 times greater among the incarcerated population compared to the non-incarcerated population. The green and yellow bars (d) and (e) indicate the period of active surveillance in prisons in Campo Grande and Dourados respectively.

### Incarceration history

We obtained permission from the Mato Grosso do Sul state prison administration agency, Agência Estadual de Administração do Sistema Penitenciário, to access a database of all prison entries, exits, and transfers within the criminal justice record system from January 1, 2005 through December 31, 2018 (**S1 Text**).

### Laboratory diagnosis and culture

All sputum specimens collected in the participating laboratories were examined by microscopy or GeneXpert® MTB/RIF, decontaminated with sodium hydroxide (NaOH), and inoculated in Ogawa-Kudoh culture medium. Cultures were incubated at 37°C for up to eight weeks and checked weekly for visible colonies at the participating laboratories. Microbial species determination was made with the MPT64 protein detection-based immunochromatographic rapid test (SD Bioline Kit, Standard Diagnostics, Inc., Korea). DNA was extracted from cultured isolates with the manual CTAB (Cetyl trimethylammonium bromide) method and whole genome sequenced on an Illumina NextSeq (2 × 151-bp).

### Whole genome sequencing

We sequenced whole *M. tuberculosis* genomes from cultured isolates on an Illumina NextSeq (2 × 151-bp). Sequence data is available on the Sequence Read Archive, in BioProject PRJNA671770. We applied variant calling methods closely following those described in Menardo et al.[23] to be consistent with the methods used for molecular clock estimation (**S1 Text**).

### Phylogenetic and Bayesian evolutionary analysis

We fit a maximum likelihood tree with RAxML-ng 1.0.1 and rooted the tree on two Lineage 1 isolates from this study.[24] We clustered genomes using a 12-SNP threshold. For each cluster including three or more isolates, we fit Bayesian trees with BEAST 2.6.2[25] with a strict clock, constant coalescent population size model using tuberculosis notification dates to calibrate tips (**S1 Text**).

### Transmission inference

*M. tuberculosis* phylogenetic trees represent patterns of evolutionary relatedness between the consensus bacterial genomes sampled from different individuals. Because most outbreaks are incompletely sampled and individuals may be infected with diverse populations of *M. tuberculosis*, phylogenies do not represent the underlying history of transmission.[21] We used TransPhylo[21] to infer transmission linkages—including unsampled hosts—that were consistent with the underlying timed phylogenies and epidemiology of tuberculosis (i.e. generation time and sampling intervals). TransPhylo samples transmission trees that are consistent with timed phylogenetic trees and epidemiological priors governing the distribution of likely *M. tuberculosis* generation times (time between subsequent infections) and sampling time (time from an individual’s infection to diagnosis) (**S1 Text**).

We then combined transmission trees with incarceration histories to identify spillover events from prisons to surrounding communities. We defined spillover events as transmission that occurred from an individual with a history of current or former incarceration to the community, including individuals with no incarceration history and prior incarceration. We considered transmission probabilities >50% as evidence of spillover and included spillover events no more than two degrees removed from prisons. TransPhylo infers transmission linkages including transmission to and from unsampled hosts and thus does not require that all transmission events be observed. Therefore, we are able to quantify the minimum number of transmission events, but not estimate the total role of spillover on community transmission.

## Results

### Tuberculosis and incarceration trends in Mato Grosso do Sul

Brazil’s incarcerated population grew by 107% (from 361,402 to 748,009) from 2005 to 2019, an increase closely paralleled in the Central West state of Mato Grosso do Sul, where the incarcerated population grew by 115% (7,891 to 16,976) over the same period (Fig. 1a-c). In 2019, prisons in the state were at 203% occupancy. While the state’s notification rate of new and retreatment tuberculosis in the general population (38 per 100,000) is similar to Brazil’s national tuberculosis notification rate (40 per 100,000) in 2019,[26] the notification rate within prisons was 43 times as high (1,666 per 100,000), again similar to Brazil’s national notification rate within prisons (1,596 per 100,000) in 2019 (Fig. 1d,e).

### Population-based genomic surveillance across Mato Grosso do Sul state

From 2014 to 2019, 3,491 new and retreatment cases of tuberculosis were reported in the Campo Grande and Dourados, the two largest cities in Mato Grosso do Sul, Brazil. Of these, 1,249 had positive cultures and we sequenced 787 *M. tuberculosis* isolates. We sequenced additional isolates from three other cities in the state and from earlier years in Dourados for a total of 1,090 isolates from unique tuberculosis notifications (Fig. 1b). Whole genome sequences (WGS) for a total of 1,043 isolates met our coverage and quality criteria (Methods). We excluded 10% (108 of 1043) of isolates with mixed infection, resulting in 935 high-quality genomes from 935 unique tuberculosis episodes from 918 individuals.

Of the 935 isolates in our analyses, 50% (465 of 935) were from patients who were incarcerated at the time of tuberculosis notification; 16% (150 of 935) were formerly incarcerated; and 34% (320 of 935) of the study population did not have an incarceration history. Among those who did not have an incarceration history, 32 people reported contact with incarcerated individuals. Additional population characteristics are in Table E1. Isolates were largely from *M. tuberculosis* lineage 4 (932 of 935) and predominantly fell into three sub-lineages: lineage 4.1, 4.3, and 4.4 (Fig. 2). We focus subsequent analyses of transmission on lineage 4. Overall, we found a low prevalence of antibiotic resistance across isolates, with 93.7% of isolates susceptible to all antibiotics (Fig. 2).

**Fig 2.**
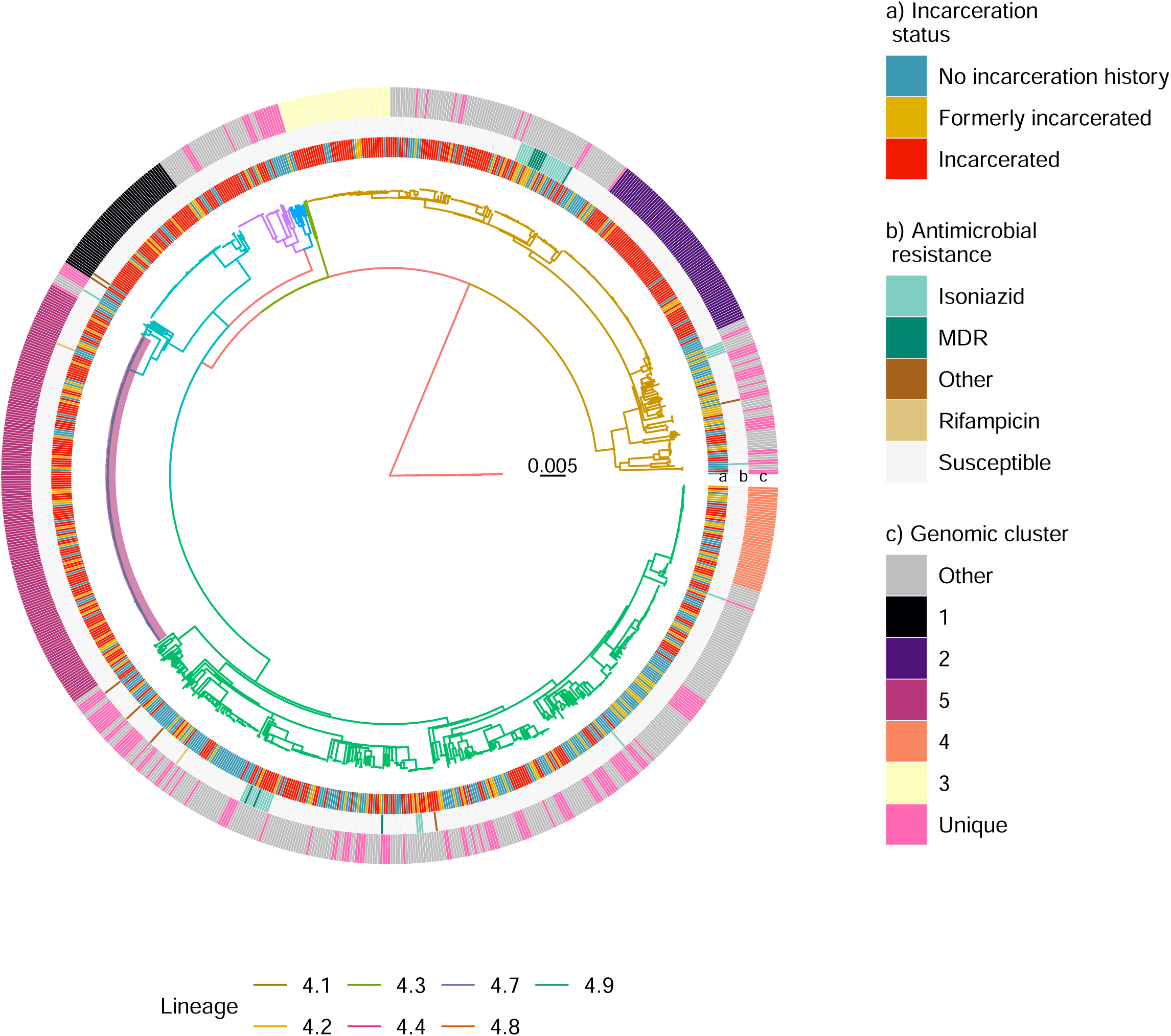
*M. tuberculosis* isolates from incarcerated and non-incarcerated people are closely related in Mato Grosso do Sul, Brazil. A maximum likelihood phylogeny of 932 tuberculosis isolates from Lineage 4 inferred from a multiple sequence alignment of 19,753 SNPs and rooted on two Lineage 1 isolates from this study. Branch lengths are in units of substitutions per site. Branches are colored by sub-lineage. From the inside, rings are colored by incarceration status, antimicrobial resistance prediction, and genomic cluster (Methods). “Other” resistance indicates resistance to at least one antibiotic other than isoniazid or rifampicin. The clade containing the largest predicted genomic cluster (Cluster 5), including 170 isolates, is highlighted in purple.

### Phylogenetic structure of *M. tuberculosis* from prisons and the community

In a maximum likelihood phylogeny (Fig. 2), *M. tuberculosis* isolates sampled from incarcerated and non-incarcerated people are distributed across the tree and do not form distinct clades, indicating recent shared evolutionary history of isolates sampled from prisons and the community. Terminal branch lengths, a proxy for the degree of recent transmission,[27] were significantly longer for isolates from people with no incarceration history at the time of tuberculosis notification (9.4 × 10^−4^ substitutions/site) compared to isolates from incarcerated individuals (2.2 × 10^−4^; p < 0.001) or formerly incarcerated individuals (3.8 × 10^−4^; p=0.019), suggesting that individuals with an incarceration history were more likely to be recently infected, although this could alternatively reflect a higher rate of genomic sampling (Fig. S1; permutation test in **S1 Text**).

We tested for phylogenetic signal in incarceration status of sampled tuberculosis patients— whether incarceration status was distributed randomly across the tree—with Fritz’s D[28], a test “clumping” in observed traits. The *D* statistic is equal to 1 if the trait is distributed randomly across tree tips with respect to the phylogeny and 0 if the trait is consistent with a pattern of clumping consistent with Brownian motion of the trait along the phylogenetic tree.

We predicted that if prison and community epidemics were distinct, incarceration status would exhibit an extremely clumped pattern on the *M. tuberculosis* phylogeny, with separate clades including samples from incarcerated people and community members. Conversely, if transmission between prisons and the community occurred just as frequently as transmission within prisons, there would be a random pattern of incarceration status across the phylogeny. We found that incarceration status had a moderate phylogenetic signal (Fritz’s D=0.59), indicating a pattern of shared evolutionary history of isolates collected from individuals with and without an incarceration history, with some degree of “clumping” with respect to incarceration status.

### Genomic clustering of *M. tuberculosis* from prisons and the community

Patterns of genomic relatedness are often used to infer potential recent transmission; *M. tuberculosis* isolates that are more closely genetically related are hypothesized to be more likely linked through recent transmission[29]. To identify potential transmission clusters, we applied a commonly used 12-SNP threshold [29], including all isolates within the threshold distance of at least one other clustered isolate. Eighty-three percent (777 of 935) of the isolates fell into 84 genomic clusters (each including 2 to 170 isolates), providing evidence that notifications were largely attributable to recent, local transmission rather than travel-associated importation or re-activation of genetically distinct latent infections (Fig. 2). Isolates from incarcerated people were more frequently clustered (93.3%, 434/465), than those from formerly incarcerated (86.0%, 129 of 150; p <0.0001), or never incarcerated people (66.9%, 214 of 320; p <0.0001), again suggesting more recent transmission within prisons.

We predicted that if prison and community-associated epidemics were distinct, isolates from the community would be most closely related to and cluster with other isolates from the community and vice versa. The majority (70.8%; 46/65) of genomic clusters including people with no incarceration history also included individuals with a recent history of incarceration. Among cases in individuals with no incarceration history, 50.6% (162/320) were in clusters that included individuals with recent incarceration history, suggesting that transmission chains often span prisons and communities.

We found a similar pattern of clustering of isolates from community members and individuals with an incarceration history when using an alternative 5-SNP threshold for genomic clustering[30]. With this threshold, of the 80 clusters, 56 included isolates from people with no incarceration history and 78.6% (44 of 56) also included isolates from people with an incarceration history. Among all tuberculosis cases occurring among individuals with no incarceration history, 46.9% (150 of 320) were in 5-SNP clusters that included individuals with incarceration history.

### A *M. tuberculosis* clone spans prisons and the community across the state

The largest potential transmission cluster, including 170 isolates sampled from September 2010 to April 2019, was distributed across the state, including cases found across state’s two largest cities, Campo Grande and Dourados, as well as the smaller cities Corumbá (on the Bolivian border) and Ponta Porã (on the Paraguayan border) (Fig. 1b, Fig. 3a). The cluster had a most recent common ancestor in 1996 (95% HPD: 1989-2003; Fig. 3a), indicating that it had circulated for approximately 23 years before the most recent samples were collected. 103 isolates were from people incarcerated at the time of diagnosis, 35 from people formerly incarcerated, and 32 from people with no incarceration history at the time of tuberculosis notification.

**Fig 3.**
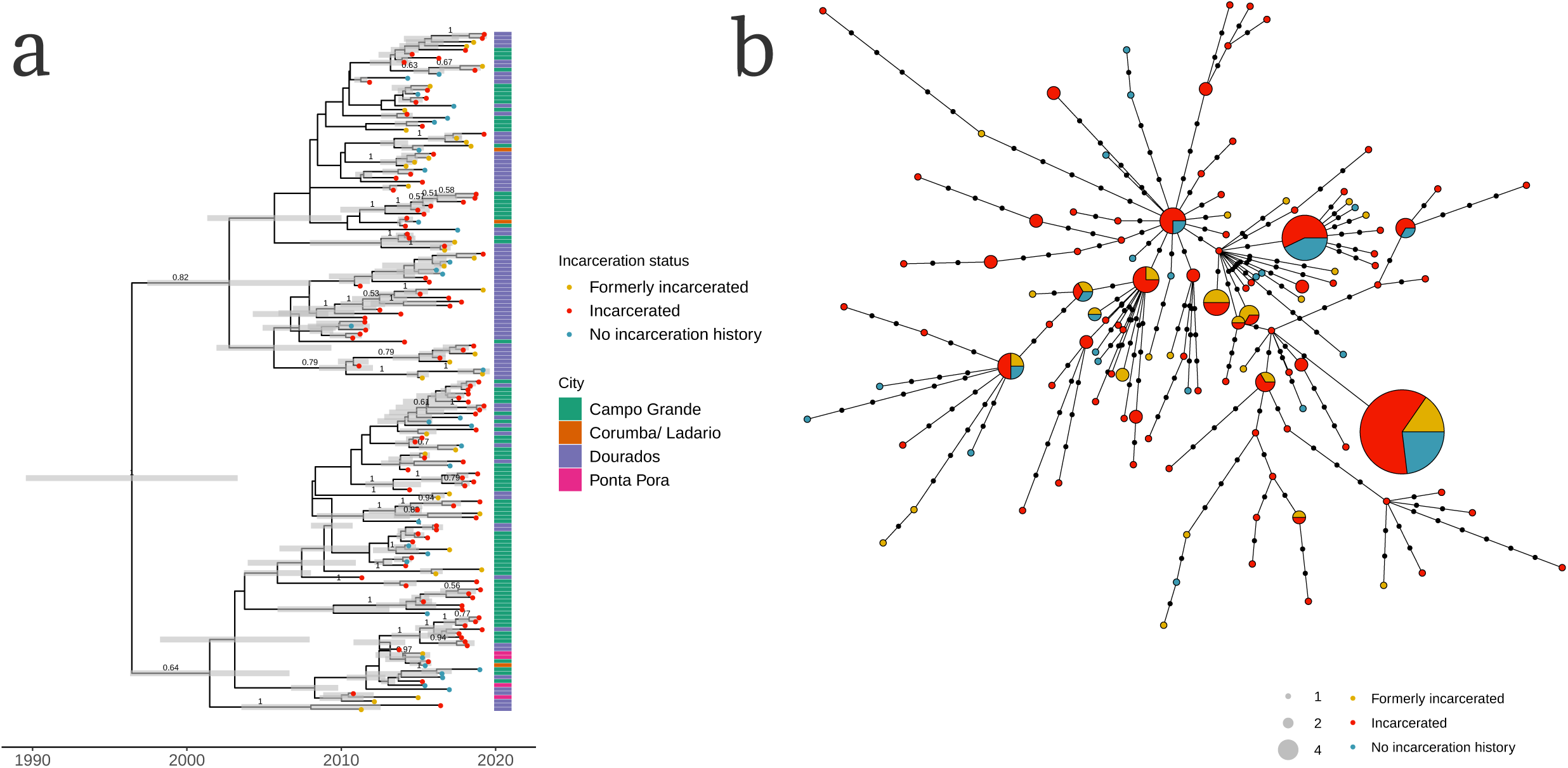
A single, recently emerged *M. tuberculosis* clone spans prisons and the community across the major cities Mato Grosso do Sul state. (a) A Bayesian time-calibrated phylogeny of the largest sampled genomic cluster of 170 isolates. The cluster emerged approximately 23 years before the most recently sampled isolate, with a most recent common ancestor in 1996 (95% HPD: 1989-2003). Tip point color indicates patient’s incarceration status at the time of tuberculosis notification. Annotation bar colors indicate city. Grey error bars indicate the 95% Bayesian highest posterior density intervals for node date. Clade posterior support values are shown on the middle of branches for clades with posterior support > 0.5. (b) A haplotype network of the single largest genomic cluster, including 170 isolates. Nodes represent unique haplotypes and are scaled to size. Points along branches indicate SNP distances between haplotypes. Node color indicates incarceration status at the time of diagnosis.

As observed in other clusters, isolates from people who are currently and formerly incarcerated were closely genetically related—and often, identical—to isolates from people who were not incarcerated at the time of notification, suggesting they are linked through recent transmission (Fig. 3b). The three largest haplotypes (identical sequences), including 14, 9, and 8 samples, were dominated by isolates from people who were incarcerated. Each haplotype also included isolates from people who were formerly incarcerated and from people with no incarceration history, indicating very recent transmission between incarcerated individuals and individuals in the community.

### Prison networks disseminate *M. tuberculosis* across space

We hypothesized that the criminal justice system’s frequent transfer of people between prisons and jails could disseminate TB across the state. We analyzed movements in the state’s incarceration database and found that the average duration of incarceration was 230 days in a prison and 25 days in a jail. The incarceration database documents 7,982 mean yearly transfers between prisons (including closed and semi-open prisons) from 2015-2018 in a prison population of 17,221 in August 2018, including frequent transfers between cities across the state (Fig. 4a).

**Fig 4.**
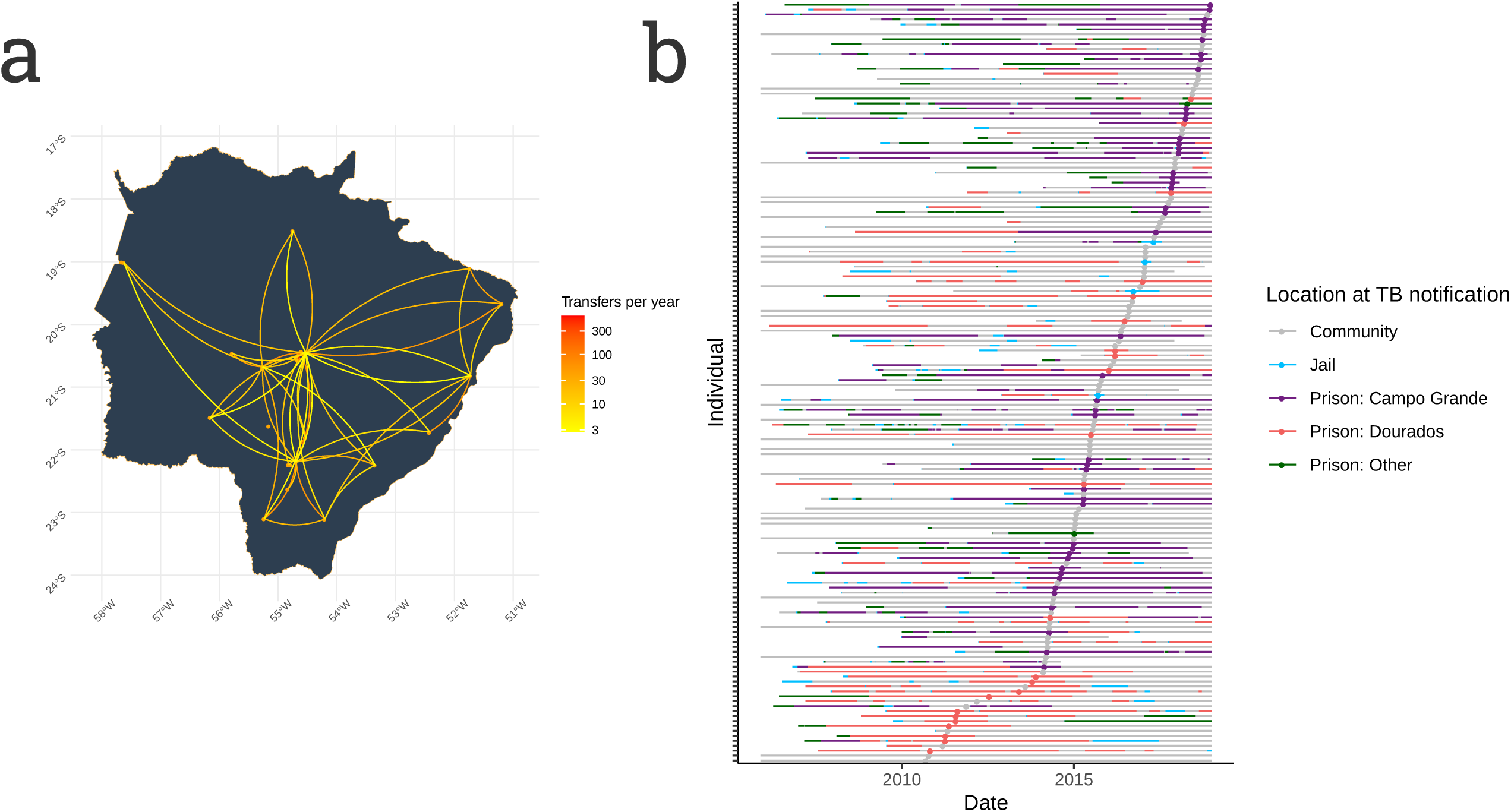
Prisons disseminate tuberculosis through the frequent movement of people between prisons. (a) Incarceration locates people within highly connected contact networks. Map of Mato Grosso do Sul state with orange points indicating the 21 prisons which most frequently transfer people between prisons. Edges are colored by the mean number of yearly transfers made between prisons from 2015-2018, including transfers in both directions. Edges were drawn between prison pairs with more than 10 transfers in the study period. Transfers between prisons within a city are not shown. (b) An *M. tuberculosis* clone (depicted in Figure 3) spreads throughout multiple prisons, jails, and the community. Each line represents an individual patient infected with an isolate in the largest genomic cluster, ordered by notification date. Line color indicates patient location, determined by matching patient names with the state incarceration database (Methods). Points indicate TB notification dates; point color indicates patient location at the time of TB notification. Individuals with notification dates after December 31, 2018 are not shown.

To investigate the role of the criminal justice system in disseminating infection, we tracked the spread of the largest sampled *M. tuberculosis* cluster (Fig. 3) across prisons in Mato Grosso do Sul (Fig. 4b). From early 2011 to 2014, sampled isolates from the cluster were concentrated in the maximum security prison in Dourados, after which it was identified within two prisons in Campo Grande and then exported to prisons and jails in other cities. The cluster was not contained within the state’s prison network; notified tuberculosis cases among community members outside of prison occurred over the duration of the clusters’ state-wide spread.

### Transmission trees reveal frequent spillover of infection

We further investigated recent transmission in the state by inferring probabilistic transmission trees representing who-infected-whom for the 84 identified *M. tuberculosis* genomic clusters with TransPhylo[21]. Because our sampling of the TB epidemic in Mato Grosso do Sul was incomplete and we did not observe the majority of transmission events resulting in sampled cases, we quantified the minimum number of recent infections attributable to prison spillover rather than the total role of spillover on the community epidemic. We identified a minimum of 16.8% (18 of 107) infections among never incarcerated individuals were directly attributable to observed spillover from prisons. Observed spillover of infection to never or formerly incarcerated individuals occurred across the state in the four largest cities sampled (Campo Grande, Dourados, Ponta Pora, and Corumba) and every year, from 2014-2019.

Together, these findings indicate that spillover of *M. tuberculosis* infection from prisons to surrounding communities is frequent. The result is that the 0.84% of the state’s population incarcerated at a given time (22,706 of 2,713,147 in 2017) or 2.6% currently or formerly incarcerated at a given time (71,703 of 2,713,147), have a disproportionate effect on TB transmission outside of prisons.

### Transmission inferences are sensitive to epidemiological priors

We conducted a sensitivity analysis to determine the effect of epidemiological priors (Table S2) on transmission inferences for the largest genomic cluster (170 isolates). Transmission probabilities inferred for pairs of individuals were closely correlated across generation time priors (Pearson’s r: 0.94 – 0.98, Fig. S3) and sampling time priors (Pearson’s r: 0.84 – 0.98, Fig. S4), although the total number of observed transmission events was strongly influenced by both priors (Figure S5). This suggests that, as expected, epidemiological priors impact the magnitude of inferred transmission probabilities rather than the identity of predicted transmission pairs. As expected, the minimum number of observed spillover events in the largest cluster were sensitive to epidemiological priors, when transmission was defined as a transmission probability greater than or equal to 50%, and ranged from 0 (short sampling and generation times) to 28 (medium generation time and very long sampling time) (Fig. S6).

## Discussion

Here, we linked intensive *M. tuberculosis* genomic surveillance to detailed individual-level incarceration histories, allowing us to reconstruct high-resolution population-wide tuberculosis transmission in Central West Brazil, a setting with an extreme disparity in tuberculosis incidence between prisons and surrounding communities. We found that *M. tuberculosis* sequences from incarcerated and non-incarcerated individuals were closely evolutionarily related and sometimes identical. Locally circulating *M. tuberculosis* clones spanned prisons and the community, and the majority of genomic clusters included individuals with and without a recent history of incarceration. In reconstructed transmission trees, we found evidence of frequent spillover of infection in all the major cities in the state and across sampling years, indicating that spillover is not the effect of a single prison but a widespread phenomenon. We found that frequent transfers of people between prisons in Mato Grosso do Sul, Brazil creates a closely connected prison contact network that likely facilitates the spread of *M. tuberculosis* between prisons and may disseminate infection risk across the state.

While alarming disparities in tuberculosis incidence between prisons and surrounding communities are well-documented,[9,13] it has been more challenging to directly link tuberculosis cases in the community to spillover from prisons. Previous studies of the broader effects of prisons on general TB epidemics have largely been associative. Here, we describe a probabilistic framework for identifying spillover events that can be applied more generally to measure the impact of high-transmission environments such as mines or hospitals on tuberculosis transmission in the general population. Our finding of frequent and widespread spillover events demonstrates the outsized effect of prison environments on community-wide *M. tuberculosis* transmission. This also suggests that the introduction of *M. tuberculosis* lineages or resistance mutations into prisons is likely to be a source of community epidemics. Our finding of the likely role of prison transfers in disseminating *M. tuberculosis* highlights an additional role that carceral systems may play in propagating infectious diseases both within highly-connected prison systems and across space.

The rapid rise in incarceration rates in many countries[31] and stark global disparities in tuberculosis incidence inside compared to outside prisons[8,9,13,32] suggests that spillover of infection may contribute to community TB transmission outside of Brazil. A study of multi-drug resistant (MDR) TB transmission found that up to 31% of MDR TB in the country of Georgia was directly or indirectly linked to prisons.[22] Our transmission inference framework is conservative in that it identifies a minimum contribution of spillover to total community transmission, and therefore, is consistent with these findings. The role of prisons and other detention centers as disease reservoirs is not unique to tuberculosis. Previous studies have identified spillover of meningococcal disease[33] and SARS-CoV-2[34] from prisons to the community. Prison transfers have also spread of SARS-CoV-2 across prisons in California, resulting in widespread outbreaks.[35]

The frequent spillover of infection from prisons to surrounding communities indicates that for TB control to be achieved in Central West Brazil, TB control efforts must focus on reducing the extraordinarily high transmission risk within prisons. Tuberculosis control programs should expand routine active screenings during[12,36] and following incarceration so that cases can be diagnosed early and linked to treatment. Individuals leaving prison are at heightened risk of tuberculosis and are a particularly vulnerable population. Preventive therapy is not provided in prisons in Brazil and many other low- and middle-income countries and could represent an effective means for tuberculosis prevention, particularly among individuals exiting prisons for community environments where the risk of reinfection is lower. More broadly, improving the unsanitary, inhumane conditions of incarceration and expanding access to primary healthcare and nutrition are critical.

Reducing this excess burden of disease will require work that extends beyond biomedical interventions. The most direct way to mitigate the excess tuberculosis risk created by prisons is for governments to rapidly reduce incarcerated populations, as advocated by the American Public Health Association, to reduce the broader harms associated with incarceration.[37] Prison releases have already begun as a means of reducing infectious disease transmission risk. [38] In an attempt to reduce the risk of COVID-19 in prisons, for example, Brazil released more than thirty thousand people by mid-2020.[39]

Our study has several limitations. First, our study highlights the uncertainty inherent in transmission inferences drawn from genomic surveillance data, which almost always constitutes an incomplete sample of population-wide infections. We addressed the incomplete sampling of *M. tuberculosis* genomes in two ways. First, we took a probabilistic transmission inference approach that accounts for uncertainty in transmission linkages and also includes unsampled individuals contributing to transmission.[21] Second, in order to collect the most comprehensive set of *M. tuberculosis* genomes possible, we combined isolates collected during active mass screening within prisons and passive routine surveillance in the community. The result was that isolates from prisons are overrepresented in our genomic collection. Because of the over-sampling within prisons, we could not directly quantify the contribution of transmission within prisons and spillover from prisons on overall recent infections in the study area. However, by taking a probabilistic transmission inference approach that accounted for uncertainty in transmission linkages as well as unsampled individuals contributing to transmission, we were able to quantify the minimum number of reconstructed spillover events consistent with inferred transmission trees. Because we cannot infer the identity or incarceration history of unsampled people who contribute to transmission, our estimates of the contribution of spillover are conservative and likely underestimate the full impact of spillover on infections in the community. However, our finding of geographically widespread spillover occurring over several years suggests that spillover is common and an important contributor to population-wide *M. tuberculosis* transmission.

Additionally, transmission trees reflect information from timed *M. tuberculosis* phylogenies and epidemiological priors and it remains difficult to distinguish genomic sampling proportion from generation time and sampling time distributions. Because we conducted transmission inference in a tuberculosis-endemic setting, rather than in a discrete outbreak, our rate of genomic sampling was lower than in some previous tuberculosis transmission studies. To reduce uncertainty in sampling proportion, we conducted transmission inference on recent subtrees for which we had sampled a greater proportion of isolates. To investigate the influence of epidemiological priors on transmission inferences, we conducted a sensitivity analysis (Table E2) and found that our conclusions about transmission linkages were largely unchanged (Supplementary Text, Figs. S3-S4), although, as expected, the number of transmission events and thus spillover events was dependent on epidemiological priors (Figs. S5-S6).

Finally, we conducted whole genome sequencing of *M. tuberculosis* isolates cultured from sputum and generated single consensus genomes from each participant, excluding within-host diversity. We also employed an existing bioinformatic approach to identify *M. tuberculosis* variants and excluded repetitive genomic regions as is common practice.[23,40] Future studies that leverage within-host *M. tuberculosis* diversity and/or include the diverse PE/PPE genes could provide greater resolution and reduce the uncertainty of transmission inferences.[41]

In this study, we present genomic evidence that spillover of tuberculosis infection risk from prisons to surrounding communities is frequent in Central West Brazil. The dramatic expansion of incarceration in recent decades has put an increasing population at extremely high risk of tuberculosis; this risk extends to surrounding communities. Reducing the excess tuberculosis transmission risk within prisons and other detention centers is therefore an urgent public health priority. Our findings additionally highlight the potential for pathogen genomic surveillance to be used to identify other key environments with a disproportionate role in pathogen transmission in order to prioritize public health interventions.

## Data Availability

Sequence data is available on the Sequence Read Archive (SRA), in BioProject PRJNA671770.

## Acknowledgments

We would like to acknowledge the contributions of AGEPEN, the Mato Grosso do Sul state criminal justice agency; LACEN, the state reference laboratory; and the Mato Grosso do Sul State Health Department. All authors had full access to all the data in the study and had final responsibility for the decision to submit.

## Notes

### Competing Interest Statement

The authors have declared no competing interest.

### Funding Statement

National Institutes of Health grant R01AI130058 (JRA) and R01 AI149620 (JRA and JC).
Brazil's National Council for Scientific and Technological Development grant 404237/2012-6 (JC).

### Author Declarations

All participants provided written consent, and this study was conducted with the approval of the Research Ethics Committee from the Federal University of Grande Dourados, Federal University of Mato Grosso do Sul and National Research Ethics Committee (CONEP) (CAAE 37237814.4.0000.5160, 2676613.3.1001.5160, and 26620619.6.0000.0021) and Stanford University Institutional Review Board (IRB-40285).

### Summary of Updates

Revised manuscript and Figures 2 and 4.

