## Supplementary Information for "The role of prisons in disseminating tuberculosis in Brazil: a genomic epidemiology study"

**This PDF file includes:**

S1 Text
Figs. S1 to S7
Tables S1 to S2

**S1 Text**

**Genomic surveillance methods**

Active screening was conducted in the three largest prisons in the state: Estabelecimento Penal "Jair Ferreira de Carvalho" (EPJFC; 2019 population 2,588) and Instituto Penal de Campo Grande (IPCG; 2019 population 1,602), a maximum and medium security prison in Campo Grande, beginning in November 2017, and Penitenciária Estadual de Dourados (PED; 2019 population 2,699), a maximum security prison in Dourados beginning in April 2018 (Fig. 1b). Screenings were conducted one to two times a year and included symptom assessment, chest radiography, sputum testing by Xpert MTB/RIF fourth-generation assay, and sputum culture. (1)

We additionally recruited patients identified by municipal health departments and who were notified in Brazil's notifiable disease registry (SINAN), from across the state, focusing on the state's two largest cities, Campo Grande and Dourados, in which active surveillance was conducted, and including additional available isolates from Corumbá, Ladário, and Ponta Porã, other major cities in the state (Fig. 1b). We additionally included participants with banked isolates from the state's diagnostic laboratory from people diagnosed between 2006 to 2013. All diagnostic tests from passive surveillance were performed in Laborátorio de Ciências da Saúde (LPCS) at the Federal University of Grande Dourados, municipalities, and the state. Patients with clinical suspicion of tuberculosis sought care at primary care providers or hospitals. Diagnostic tests were performed at the city public health diagnostic laboratories. Patients with non-tuberculous mycobacteria or without positive cultures for *M. tuberculosis* were excluded. After the result of the positive culture, a team of researchers carried out a home or prison visit to recruit participants and administer the study questionnaire. There was no incentive to study participation.

Study participants answered a structured questionnaire including information on birthplace; residential address and residential history; previous history of tuberculosis diagnosis, treatment, and treatment outcomes; potential contact with patients diagnosed with pulmonary tuberculosis; incarceration history (incarcerated at the time of diagnosis, formerly incarcerated, any contact with those incarcerated, or no contact with incarcerated people); and travel history. Additional sociodemographic and clinical data were obtained from the National Reporting System on Notifiable Diseases (SINAN). Data was managed in an electronic database (REDCap).

**Incarceration history**

We matched names from the tuberculosis registry (SINAN) with the incarceration database, SIGO, using Levenshtein string distances between names and mother's names with the R fuzzyjoin package (2) and excluded matches below a 90% matching score. We manually confirmed matches for whom there was a name match in the SIGO database but no report of current or previous incarceration in SINAN or in the patient questionnaire. We defined someone as incarcerated at the time of tuberculosis notification if they had a record of incarceration at the time of notification in SIGO, were recorded as incarcerated in SINAN, or reported current incarceration in the patient questionnaire. Brazil's national criminal law defines different "regimes" or stages of incarceration (3), which may facilitate the potential for spillover of infection. People are incarcerated under closed regimes, within prisons; semi-open regimes, in which people may work outside prisons and return at night; and open regimes, under which people serve sentences outside of prison, but are required to make periodic court appearances. (3)

We defined incarceration location using the SIGO database, which had most detailed information. If there was no location in SIGO, we used location information from SINAN or the patient questionnaire. For 15 patients who were incarcerated at the time of tuberculosis notification according to SINAN, there was no information about location of incarceration. For individuals who were reported as incarcerated at the time of tuberculosis notification in study questionnaires or SINAN but who did not have a match in SIGO, we considered as being incarcerated for one year prior to their tuberculosis notification.

We obtained permission to access the Mato Grosso do Sul state incarceration database from 2005-2018. Therefore, if participants were incarcerated outside Mato Grosso do Sul state, before 2005, or if we were unable to match their tuberculosis record with an incarceration record, we could misclassify participants as having no incarceration history. We expect that this would not substantially change our conclusions as the majority of tuberculosis disease occurs within two years of infection.

We reported the 2017 state population incarcerated in 2017 as the total number of unique individuals incarcerated in a closed prison, semi-open prison, or jail on January 1, 2017, according to SIGO. We reported the 2017 state population who were currently or formerly incarcerated in 2017 as the total number of unique individuals incarcerated in a closed prison, semi-open prison, or jail from January 1, 2012 to January 1, 2017, in order to estimate the population size of people with recent incarceration history.

**Whole genome sequencing  and bioinformatic methods**

We trimmed low-quality bases (Phred-scaled base quality < 20) and removed adapters with Trim Galore (stringency=3) (4). We used CutAdapt to further filter reads (--nextseq-trim=20  --minimum-length=20 --pair-filter=any) (5). To exclude potential contamination, we used Kraken2 (6) to taxonomically classify reads and removed reads that were not assigned to the *Mycobacterium* genus or that were assigned to a *Mycobacterium* species other than *M. tuberculosis*. We mapped reads with bwa v. 0.7.15 (7) (bwa mem) to the H37Rv reference genome (NCBI Accession: NC_000962.3) and performed local read realignment with the RealignerTargetCreator and IndelRealigner modules of GATK v3.8. We created read pileups with Samtools v1.9 and called variants for individual samples with varscan v2.4.4. As described in, (8) we called variants at positions with a minimum mapping quality of 20; minimum base quality of 20; minimum read depth of 7X; and no more than 90%, or less than 10% of reads supporting a call in the same orientation (varscan strand bias filter).

We excluded SNPs in previously defined repetitive regions (PPE and PE-PGRS genes, phages, insertion sequences and repeats longer than 50 bp). (9)  We excluded all isolates with mean coverage < 15 X and isolates with more than 50% of SNPs failing the strand bias filter, and genomes with more than 50% of SNPs with a variant allele frequency between 10% and 90%. We identified mixed *M. tuberculosis* infections with QuantTB (10) and excluded isolates classified as mixed infections. We set indels to “no calls” and constructed full-length consensus FASTA sequences from VCF files and used SNP-sites to extract a multiple alignment of internal variant sites. (11) We measured drug resistance associated mutations with MykrobePredictor v0.8.0, (12) using the '201901' database of genomic predictors of resistance. (13) We identified lineage with TBProfiler v.2.8.6. (14)

**Phylogenetic and Bayesian evolutionary analysis**

We used the R package *ape* to measure the number of pairwise site differences between samples (15) and applied a commonly used 12-SNP threshold (16) to cluster isolates potentially linked through recent transmission. Missing sites, "Ns" in the alignment, were not counted as pairwise differences.

We fit maximum likelihood trees with RAxML-ng 1.0.1. (17) and used a GTR substitution model and a Stamatakis ascertainment bias correction (18) for invariant sites in our alignment. We divided the number of invariant sites by 1000 to avoid issues created by small branch lengths. We defined nucleotide stationary frequencies as frequencies in the reference genome.

We expected lineage 4 isolates from our study to share a common substitution rate. Because it was not be possible to estimate substitution rates from small trees for our Bayesian timed tree inference, we estimated substitution rate for the largest cluster (170 tips) using a log-normal prior on substitution rate (mean = -17, s = 2), consistent with previous estimates of *M. tuberculosis* lineage 4 substitution rate (mean = $5.8 \times 10^{-8}$, standard deviation = $2.0 \times 10^{-8}$). (8) We then fit a log-normal distribution to the posterior samples of the substitution rate for the largest cluster and used this as a narrow log-normal prior on substitution rate (m = -16.5, sd=0.17) for all other clusters. We used an HKY substitution rate model and estimated base frequencies. We ran sampling chains for 100 million iterations, discarding a burn-in of 10% of samples, and confirmed convergence using effective sample size estimates (confirming all parameters had ESS > 200) in Tracer 1.7.1. (19) We used TreeAnnotator v2.6.2 to summarize posterior trees as maximum clade credibility (MCC) trees, with median node heights. We corrected for ascertainment bias by specifying the number of invariant sites in the BEAST XML file.

To test for temporal signal in the largest cluster (170 tips), we performed a date randomization test with the R package TipDatingBeast. (20) We randomized dates across all tips to generate twenty date-randomized XML files. We ran BEAST with priors as specified above, and compared the posterior distribution of substitution rate estimated from the date-randomized inputs and the real data. The 95% Bayesian credible interval of substitution rate estimated from the real data does not overlap that of the substitution rates estimated with randomized dates, evidence of a temporal signal in the largest cluster (Fig. S7).

To test if terminal branch lengths differed significantly between isolates from participants with different incarceration histories, we conducted a permutation test. We randomly shuffled participant incarceration status across tips 1000 times and determined the mean difference in terminal branch lengths between groups. We used the distribution of mean terminal branch length differences to quantify the probability that observed differences in terminal branch lengths were different from the random expectation.

We tested for phylogenetic signal or "clumping" in incarceration status of sampled tuberculosis patients with Fritz's D (21), with the R package caper (22). Calues of *D* can are not restricted to the range of 0 or 1; values larger than 1 indicate the distribution is overdispersed with respect to the underlying phylogeny and less than 0 indicate extreme clumping (21). Fritz's D tests for phylogenetic signal or "clumping" in binary traits; we considered location of each study participant at the time of tuberculosis notification as inside prison or outside (including those participants formerly incarcerated).

**Transmission tree inference**

To focus on densely sampled trees that would not be dominated by unsampled hosts, we sliced phylogenies at 2012 in order to generate subtrees with a most recent common ancestor of 2012 or later for transmission inference, including 122 subtrees each with 2 to 23 tips and comprising 56% of isolates (528/935). Many transmission events are unobserved because of incomplete sampling; therefore, the sum of transmission probabilities to any individual is often less than one.

We fit transmission trees for all subtrees within the same cluster using the multiple transmission tree inference algorithm in TransPhylo in which parameters for the offspring distribution and sampling proportion are shared across trees. For both generation time and sampling time, we set gamma distribution shape and scale priors of 2.35 and 0.54, corresponding to a mean of 1.27 years and within the range of priors applied in previous studies.(23–25) We set beta distribution shape and scale priors of 1 and 1 for genomic sampling proportion, the default priors. We fixed the within-host population size (Neg) as 1.48 as done previously(24) and set the time at which the epidemiological process stops to be one year later than the most recently sampled tree tip. We discarded 20% of samples as burn-in and ran chains for one million iterations. We additionally tested the sensitivity of transmission probabilities to epidemiological priors on generation time and sampling time (below). We sampled from the posterior distribution of transmission trees, to estimate who acquired infection from whom matrices, W, where $W_{i,j}$ is the posterior probability of transmission from individual $j$ to individual $i$.

**Sensitivity analysis**

To assess the sensitivity of transmission inferences to epidemiological priors, we conducted two sensitivity analyses for the largest genomic cluster (including 170 isolates). First, we fit transmission trees under a range of priors for generation time and sampling time (Table S2). We examined correlations between pairwise transmission probabilities inferred when varying the prior on sampling time, while holding generation time constant, and vice versa (Figs. S3 and S4). We also examined the sensitivity of total observed transmissions to generation time and sampling time priors (Fig. S5). Finally, we examined the sensitivity of the population-wide WAIFW matrix to epidemiological priors (Fig. S6).

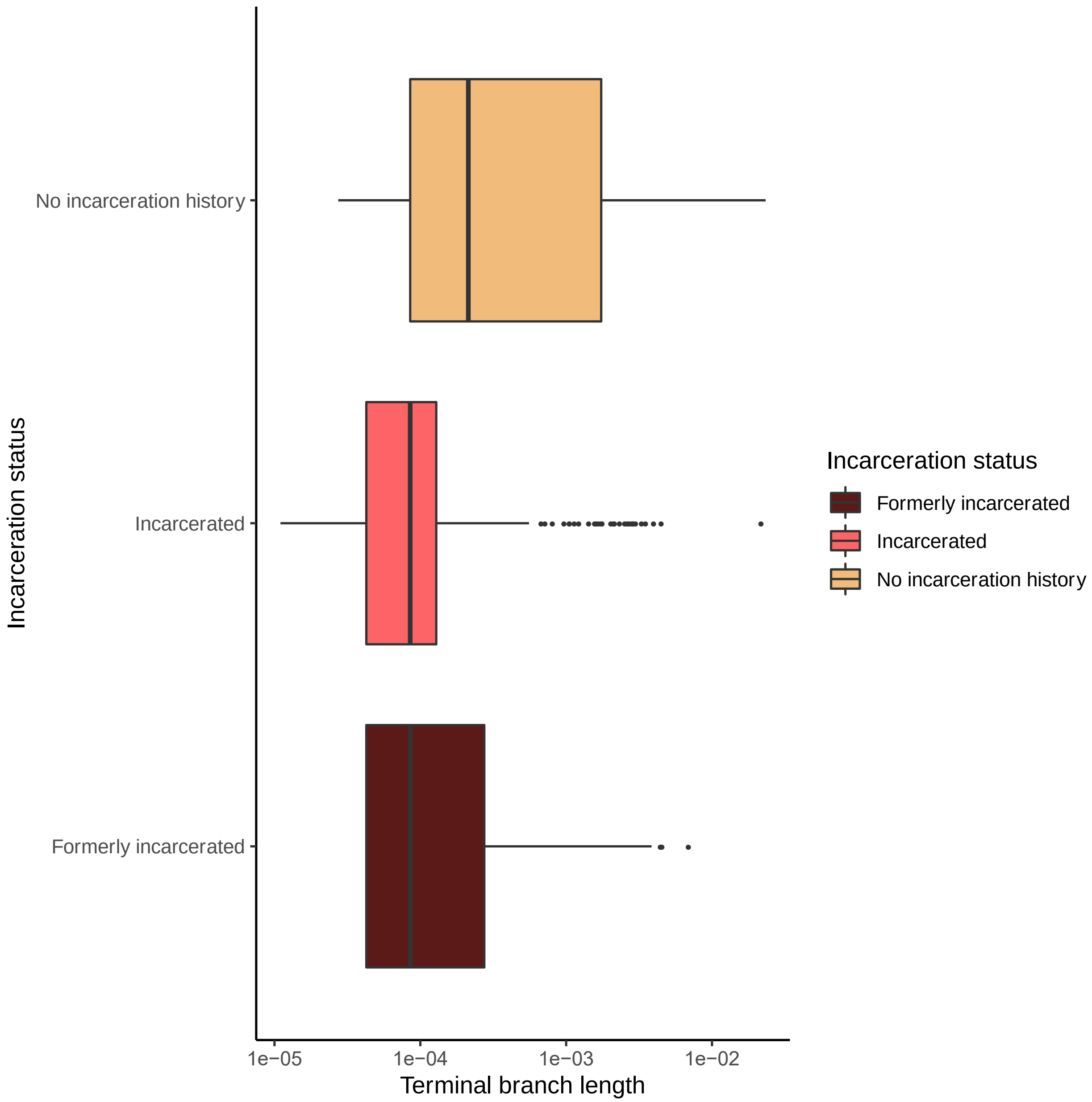


**Figure S1. Phylogenetic terminal branch lengths stratified by incarceration status at the time of tuberculosis notification.** Terminal branch lengths extracted from the RAxML phylogeny. We set near-zero branch lengths ($10^{-9}$ substitutions per site) to zero.

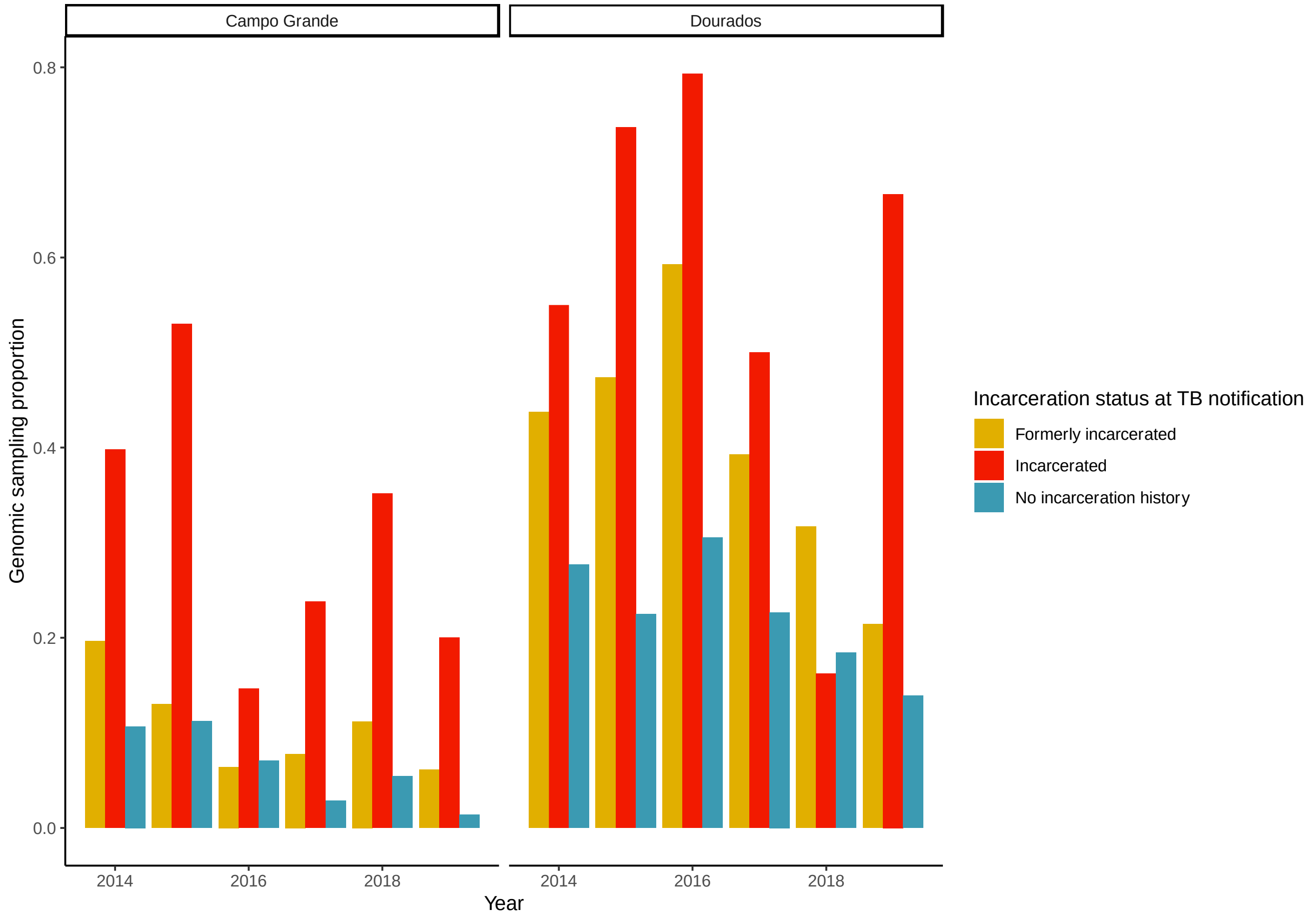


**Figure S2. Genomic sampling proportion in Campo Grande and Dourados, Brazil, 2014-2019.** To correct transmission inferences for potential over-sampling of specific populations, we determined the genomic sampling proportion, which we defined as the number of sequenced *M. tuberculosis* genomes passing all quality filters for each population divided by the number of tuberculosis notifications for each population from 2014-2019 in Campo Grande and Dourados, the two major cities in our prospective study.

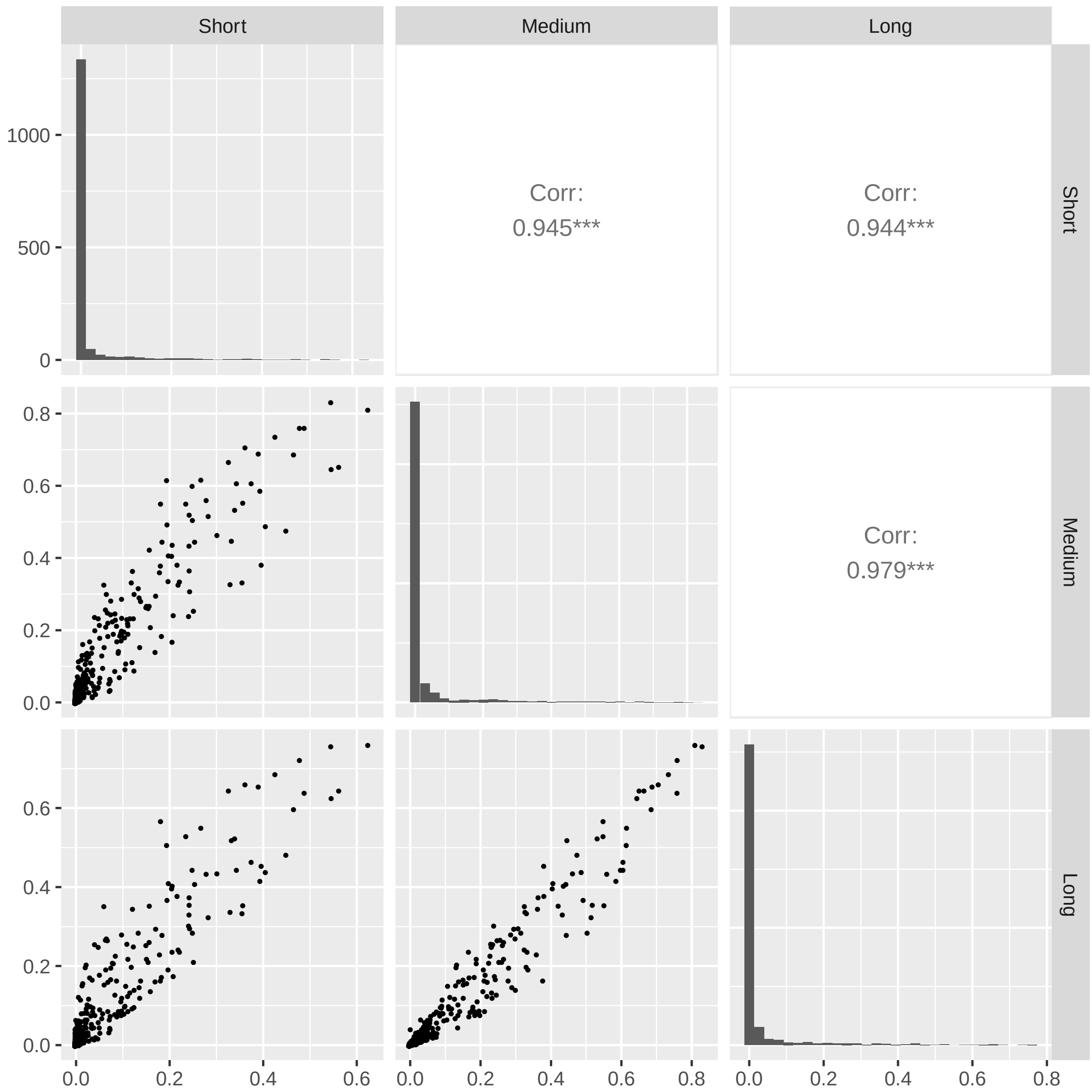


**Figure S3. Correlation of transmission probabilities inferred with different generation time priors.** Transmission probabilities inferred between pairs of isolates in the largest cluster (170 isolates) across different generation time priors (lower triangle), histograms of transmission probabilities (diagonal), and Pearson correlation coefficients for transmission probabilities inferred with different priors (upper triangle). Priors are in Table S2. Sampling time was held constant to the “medium” sampling time (Table S2).

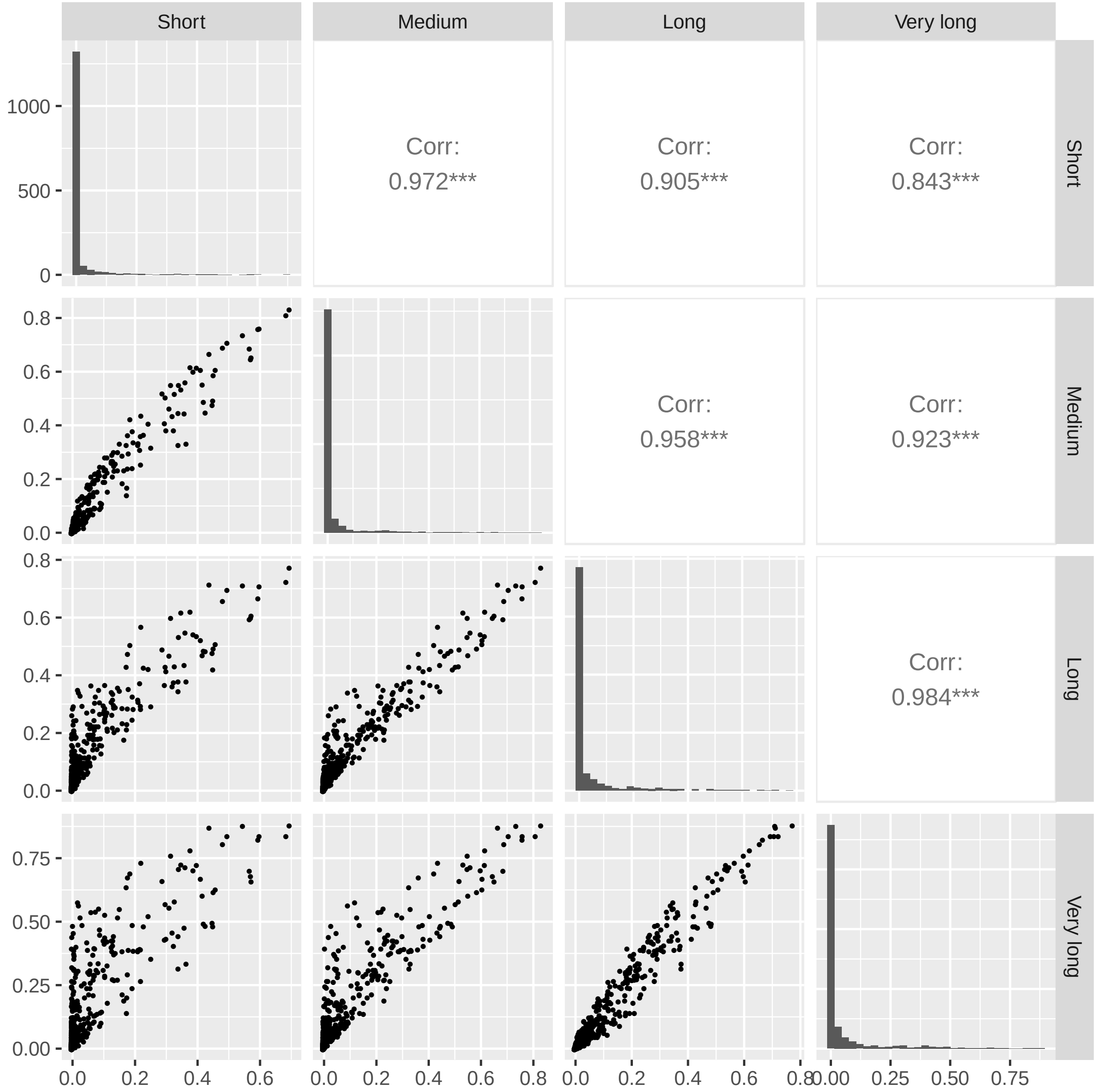


**Figure S4. Correlation of transmission probabilities inferred with different sampling time priors.** Transmission probabilities inferred between pairs of isolates in the largest cluster (170 isolates) across different sampling time priors (lower triangle), histograms of transmission probabilities (diagonal), and Pearson correlation coefficients for transmission probabilities inferred with different priors (upper triangle). Priors are in Table S2. Generation time was held constant to the "medium" sampling time (Table S2).

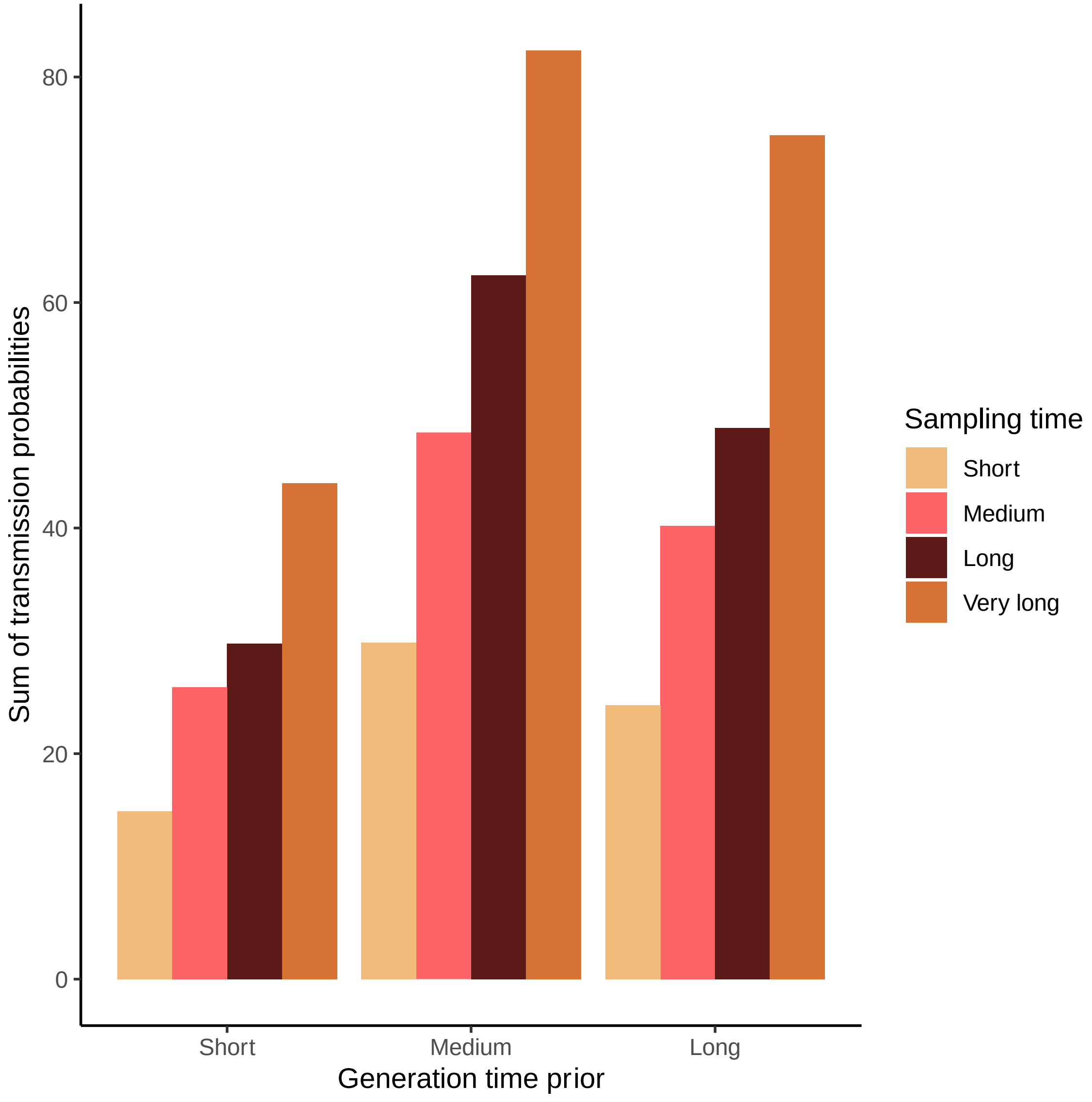


**Figure S5. Sensitivity of total observed transmissions across generation time and sampling time priors.** The sum of transmission probabilities between all potential pairs in the largest cluster (170 isolates) inferred with different generation time (x-axis) and sampling time (bar colors) priors. Priors are in Table S2. All parameters were estimated as in Figs. S3-S4.

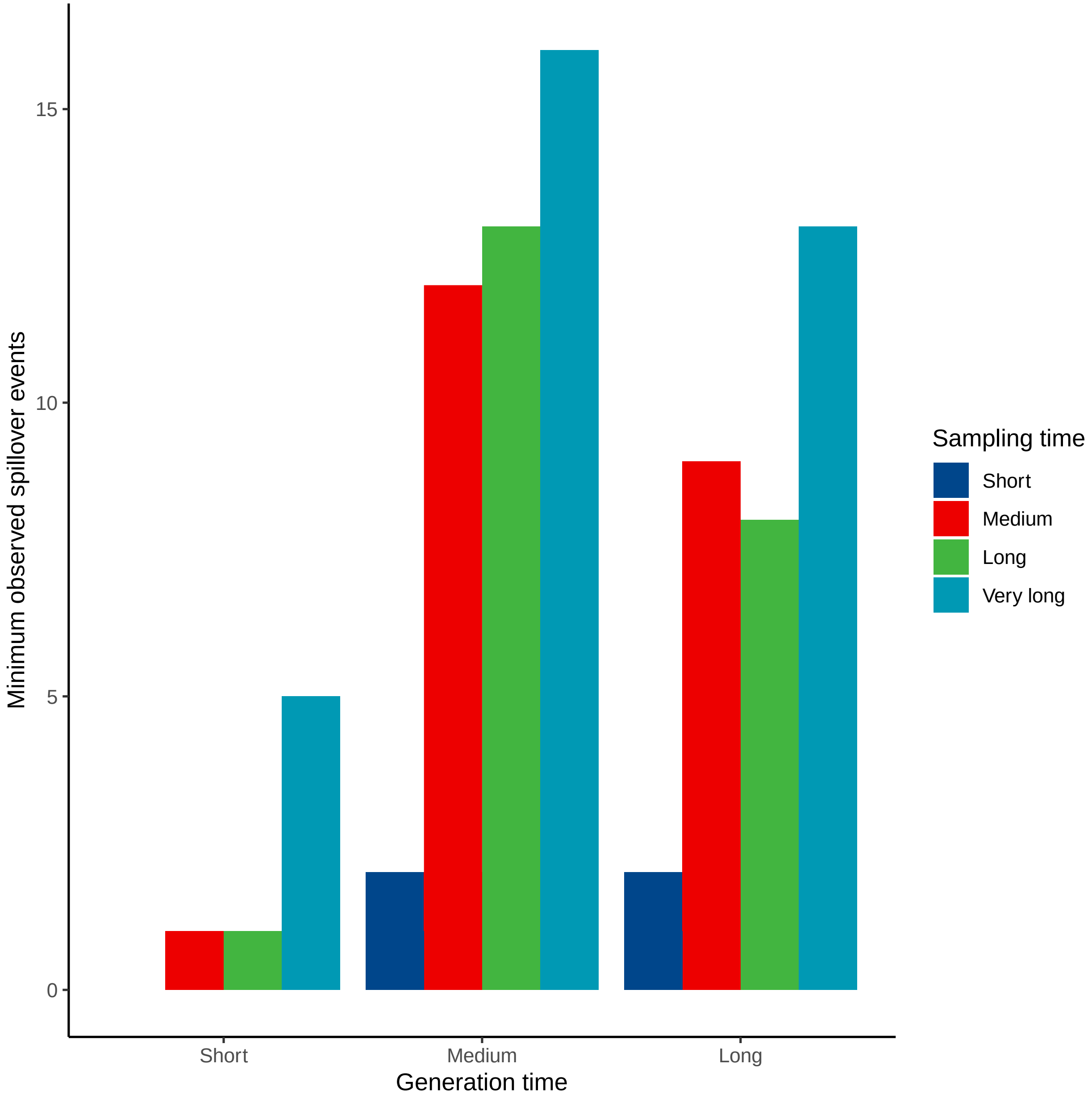


**Figure S6. Sensitivity of the minimum observed spillover events across generation time and sampling time priors.** The minimum number of spillover events, defined as transmission probabilities of >50% from a currently or formerly incarcerated individual to an individual with no incarceration history, identified in transmission reconstruction in the largest genomic cluster (170 isolates). Spillover events were inferred with different generation time (x-axis) and sampling time (bar colors) priors. The number of reconstructed spillover events represent a minimum bound on total number of unique spillover events because of incomplete epidemic sampling. Priors are in Table S2. All parameters were estimated as in Figs. S3-S4.

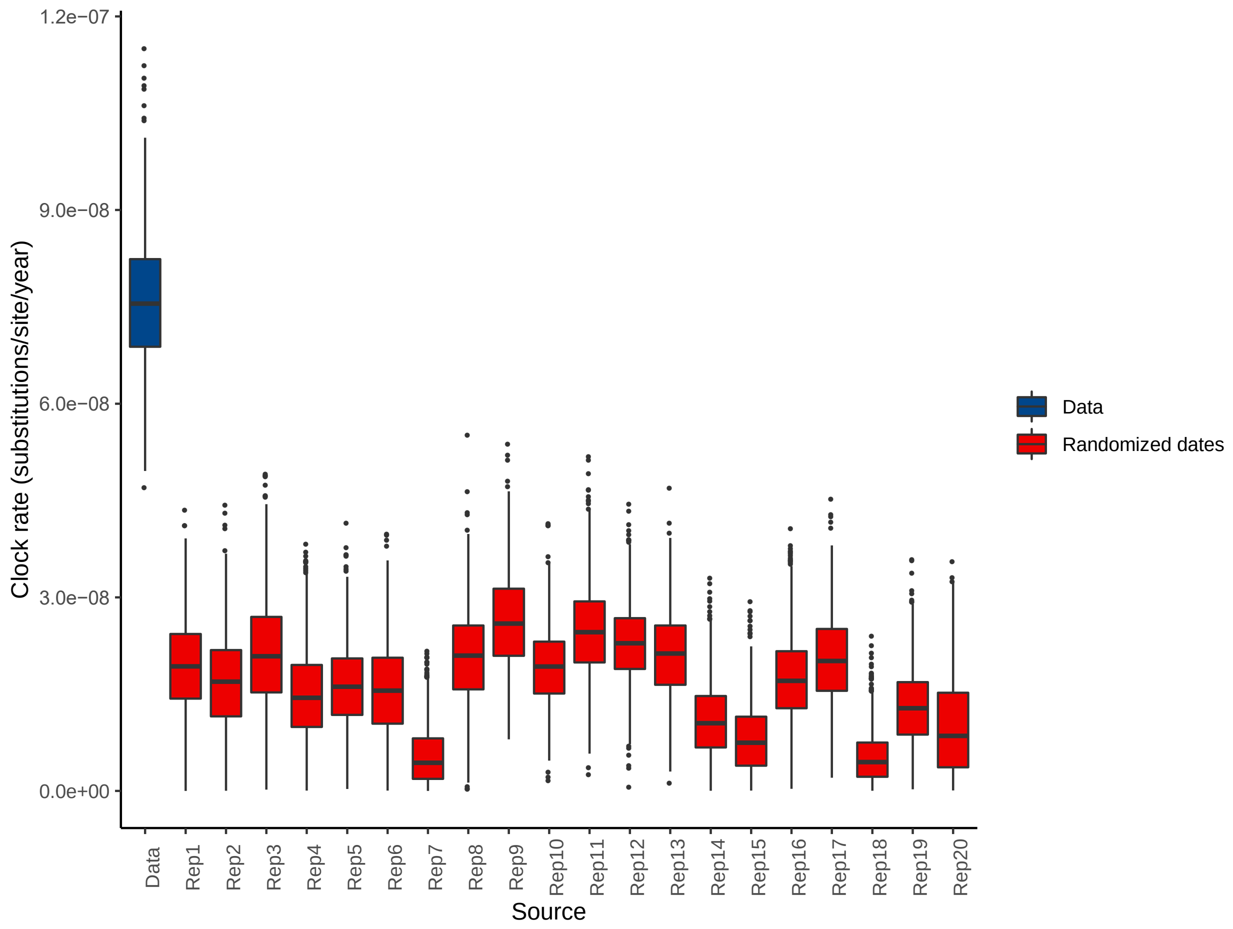


**Figure S7. Date randomization plot.** We conducted a date randomization test to determine if the largest genomic cluster in our sample (170 isolates) was a measurably evolving population. Box plots show posterior distribution of clock rate estimated from the time-stamped SNP alignment (blue) and from twenty date-randomized SNP alignments (red). The 95% Bayesian credible interval of the time-stamped samples does not overlap that of date randomized tips, supporting a measurable molecular clock for the largest cluster.

| Variable | Levels | Total |
|---|---|---|
| **Sex** | Male | 788 (84.3) |
| | Female | 146 (15.6) |
| | Unknown | 1 (0.1) |
| **Age** | mean (sd) | 34.9 (13.1) |
| | Unknown | 12 |
| **City** | Campo Grande | 400 (42.8) |
| | Dourados | 405 (43.3) |
| | Corumbá | 102 (10.9) |
| | Ponta Porã | 24 (2.6) |
| | Ladário | 2 (0.2) |
| | Camapuã | 1 (0.1) |
| | Outside Mato Grosso do Sul | 1 (0.1) |
| **Year** | 2006 | 1 (0.1) |
| | 2009 | 5 (0.5) |
| | 2010 | 21 (2.2) |
| | 2011 | 25 (2.7) |
| | 2012 | 26 (2.8) |
| | 2013 | 32 (3.4) |
| | 2014 | 163 (17.4) |
| | 2015 | 188 (20.1) |
| | 2016 | 148 (15.8) |
| | 2017 | 117 (12.5) |
| | 2018 | 165 (17.6) |
| | 2019 | 44 (4.7) |
| **Incarceration status** | No incarceration history | 320 (34.2) |
| | Formerly incarcerated | 150 (16.0) |
| | Incarcerated | 465 (49.7) |

**Table S1. Demographic characteristics of 935 participants included in genomic analysis.**

| Generation time | | | | Sampling time | | | |
|---|---|---|---|---|---|---|---|
| **Prior** | **Shape** | **Scale** | **Mean** | **Prior** | **Shape** | **Scale** | **Mean** |
| **Short** | 1.30 | 0.54 | 0.70 | **Short** | 1.50 | 0.54 | 0.81 |
| **Short** | 1.30 | 0.54 | 0.70 | **Medium** | 2.35 | 0.54 | 1.27 |
| **Short** | 1.30 | 0.54 | 0.70 | **Long** | 1.50 | 1.74 | 2.61 |
| **Short** | 1.30 | 0.54 | 0.70 | **Very long** | 2.50 | 1.20 | 3.00 |
| **Medium** | 2.35 | 0.54 | 1.27 | **Short** | 1.50 | 0.54 | 0.81 |
| **Medium** | 2.35 | 0.54 | 1.27 | **Medium** | 2.35 | 0.54 | 1.27 |
| **Medium** | 2.35 | 0.54 | 1.27 | **Long** | 1.50 | 1.74 | 2.61 |
| **Medium** | 2.35 | 0.54 | 1.27 | **Very long** | 2.50 | 1.20 | 3.00 |
| **Long** | 1.30 | 2.50 | 3.25 | **Short** | 1.50 | 0.54 | 0.81 |
| **Long** | 1.30 | 2.50 | 3.25 | **Medium** | 2.35 | 0.54 | 1.27 |
| **Long** | 1.30 | 2.50 | 3.25 | **Long** | 1.50 | 1.74 | 2.61 |
| **Long** | 1.30 | 2.50 | 3.25 | **Very long** | 2.50 | 1.20 | 3.00 |

**Table S2. Transmission inference priors for sensitivity analysis.** We conducted sensitivity analysis with three different generation time (time between subsequent infections) and four different sampling time (time between infection and genomic sampling) priors. Priors are defined by the shape and scale parameters of the Gamma distribution, the table also includes the mean time in years. The main text describes results using the “medium” generation time and sampling time priors.
